# Demographic disparities in clinical outcomes of COVID-19: data from a statewide cohort in South Carolina

**DOI:** 10.1101/2021.05.19.21257489

**Authors:** Xueying Yang, Jiajia Zhang, Shujie Chen, Bankole Olatosi, Larisa Bruner, Abdoulaye Diedhiou, Cheryl Scott, Ali Mansaray, Sharon Weissman, Xiaoming Li

## Abstract

**Background:** Current literature examining the clinical characteristics of COVID-19 patients under-represent COVID-19 cases who were either asymptomatic or had a mild illness.

**Objective:** To generate a state-level description and examine the demographic disparities of clinical outcomes of COVID-19.

**Design:** Statewide population-based cohort study

**Setting:** COVID-19 surveillance facilities in South Carolina

**Patients:** Adults COVID-19 cases reported to the SC DHEC by Case Report Form during March 04–December 31, 2020

**Measurements:** The primary predictors were socio-demographic characteristics. The outcomes were COVID-19 disease severity, hospitalization, and mortality, which collected from the standardized CRF.

**Results:** Among a total of 280,177 COVID-19 cases, 5.2% (14,451) were hospitalized and 1.9% (5,308) died. Individuals who were older, male gender, Blacks, Hispanic or Latino, and residing in small towns had higher odds for hospitalization and death from COVID-19 (Ps<0.0001). Regarding disease severity, 144,157 (51.5%) were asymptomatic, while 34.4% and 14.2% had mild and moderate/severe symptoms, respectively. Older individuals (OR: 1.14, 95%CI: 1.11, 1.18), Hispanic or Latino (OR: 2.07; 95%CI: 1.96, 2.18), and people residing in small towns (OR: 1.15; 95%CI: 1.08, 1.23) had higher odds of experiencing moderate/severe symptoms, while male and Asian (vs Whites) patients had lower odds of experiencing moderate/severe symptoms.

**Limitations:** Potential misclassification of outcomes due to missing data; other variables were not evaluated, such as comorbidities.

**Conclusion:** As the first statewide population-based study using data from multiple healthcare systems with a long follow-up period in the US, we provide a more generalizable picture of COVID-19 symptoms and clinical outcomes. The findings from this study reinforce the fact that rural residence, racial and ethnic social determinants of health, unfortunately, remain predictors of poor health outcomes for COVID-19 patients.

## Introduction

Since the first confirmed case of coronavirus disease 2019 (COVID-19), caused by severe acute respiratory syndrome coronavirus (SARS-CoV-2) in the United States (US) on January 21, 2020, outbreaks of COVID-19 have surged quickly. The US is among the countries hardest hit by the pandemic(1). As of May 19, 2021, there were over 33 million COVID-19 cases with 587,640 deaths in the US. South Carolina, a predominately rural state with significant health care shortage regions, has reported 461,681 COVID-19 cases and 9,122 deaths as of March 29(2), 2021 and ranked 12th across the nation in number of COVID-19 case(3).

The clinical spectrum of SARS-CoV-2 infection ranges from asymptomatic to life-threatening and death. Studies show presenting symptoms plays a fundamental role to inform the disease severity after COVID-19 diagnosis. Based on existing research, most individuals infected with SARS-CoV-2 are asymptomatic (around 40%-45%(4)) or experience mild to moderate symptoms(1). About 14% of all cases become severe, and 5% critical(5). Ongoing research continues to investigate clinical outcomes of COVID-19 patients in the US. However, a large heterogeneity existed between these studies because of differences in clinical settings, sample selection methods and statistical plans, which limit the generalizability of these findings.

Characteristics and clinical outcomes of COVID-19 patients has been frequently reported in existing literature, but the data are not necessarily representative of the full spectrum of disease. Most of these studies revealed an increasing evidence that some racial and ethnic minority groups (e.g., Black, Hispanic or Latino) are overrepresented in COVID-19 case(6-10) and reported a disproportionate burden of hospitalizations, but not necessarily critical illness or death(11), in these groups(12, 13). Despite an increasing body of US studies investigating clinical outcomes of COVID-19 patients, including hospitalization(7-9, 11, 14-18), mortality(9, 11, 14-17, 19), and intensive care unit (ICU) admission(17, 19), several knowledge gaps persist in existing research. First, the majority of patient samples were restricted to hospitalized COVID-19 cases or identified from a single health care system, which might be less representative to the majority of outpatient COVID-19 population who are either asymptomatic or had mild illness. Second, most of these studies collected data from a city-level or hospital level with relatively small sample sizes (305 to 78,323) and short observational study period (1-2 months), further restricting the generalization of the findings to all COVD-19 populations. Third, presenting symptoms of COVID-19 patients, particularly non-hospitalized cases, were not previously extensively investigated. We proposed to address these gaps by using data from a population-based statewide cohort which included all adult confirmed and probable COVID-19 cases in SC between March 04, 2020 and December 31, 2021. The present study analyzed sociodemographic characteristics, disease severity and clinical outcomes of COVID-19 patients, including hospitalization and mortality.

## Methods

### Data source

Data for this study were derived from the SC statewide Case Report Form (CRF) (“Human Infection with 2019 Novel Coronavirus Case Report Form”) for SARS-CoV-2 infection issued by SC DHEC. The CRF contains information about lab-confirmed and probable cases of COVID-19, including the case classification and identification, hospitalization, ICU and death information, case demographics, clinical course, symptoms, past medical history and social history. A total of 280,177 adult COVID-19 patients (age≥18 years old) who meet the standardized surveillance case definition for COVID-19(20) between March 04, 2020 and December 31, 2020 were included in the current study. SC Law (**44-29-10**) and Regulations (**61-20**) require mandatory reporting of COVID-19 to DHEC(21). Where available, reports must include information about diagnoses, and results of specific diagnostic tests. The sources of data include clinician reporting, laboratory reporting, reporting by other entities (e.g., hospitals, veterinarians, pharmacies, poison centers), death certificates, hospital discharge or outpatient records(20). The criteria of case ascertainment was described in the standardized surveillance case definition of COVID-19(20). The study protocol received approval from the institutional review board in University of South Carolina and relevant SC state agencies.

### Measures

#### Case demographics

Information on social demographics included age (e.g., 40-49, 50-59 years old), gender (e.g., female, male, transgender), race (e.g., White, Black, Asian), ethnicity (e.g., Hispanic/Latino, non-Hispanic/Latino), and residential status. Residential status was defined according to the Rural-Urban Commuting Area (RUCA) codes, as urban areas (i.e., urban focused) or rural areas (i.e., large rural city/towns or small and isolated rural towns focused) (22). Specifically, counties were grouped into micropolitan, metropolitan, rural and small-town areas.

#### Clinical course and symptoms

Clinical course information included symptom category during onset of illness (i.e., symptomatic, asymptomatic, unknown), and development of pneumonia and acute respiratory distress syndrome (ARDS). For symptomatic patients, the CRF documented specific symptoms that were experienced during the illness, such as fever, chills, muscle aches, runny nose, sore throat, new olfactory and taste disorders, headache, fatigue, cough, difficulty breathing, nausea or vomiting, abdominal pain, and diarrhea (Figure 1).

**Figure 1.**
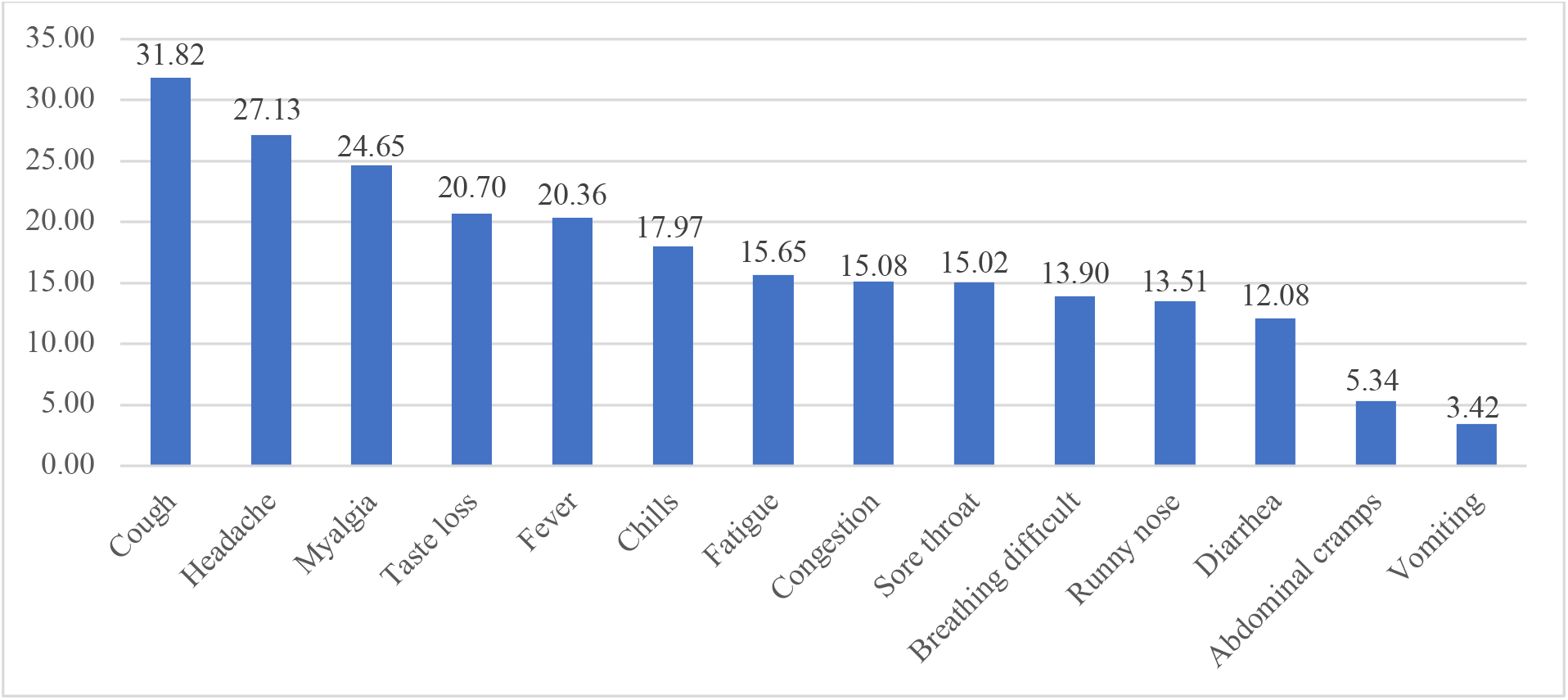
Percentages of symptoms among COVID-19 patients in South Carolina (n=280,177)

#### Outcomes

We analyzed three distinct outcomes: disease severity, hospitalization, and mortality of COVID- 19. Individuals were asked to indicate any of the symptoms (e.g., fever, headache, fatigue) specified on the CRF form. Each symptom had three responses, i.e., ‘Yes’, ‘No’, ‘Unknown’. Based on different presenting symptoms of COVID-19 patients, disease severity was categorized into three groups. Specifically, COVID-19 patients with no symptoms were categorized as asymptomatic; individuals who have any of the various mild signs and symptoms of COVID-19 (e.g., fever, cough, sore throat, malaise, headache, muscle pain, nausea, vomiting, diarrhea, loss of taste and smell) were categorized as mild; whereas COVID-19 patients with difficulty breathing or developed pneumonia or ARDS were categorized as moderate/severe. Since we do not have other clinical indicators to separate moderate with severe illness, we combined them together. In the CRF, hospitalization was measured with one question, i.e., “Was the patient hospitalized?” with the responses categorized as ‘Yes’, ‘No’, and ‘Unknown’. For patients with no response to this question, we also treated as unknown. Then we dichotomized the hospitalization status as 1 if the response is ‘Yes’ else it was defined as 0, indicating no hospital admission. Similarly, death was measured using the question, i.e., “Did the patient die as a result of this illness?”, with the response categories as ‘Yes”, “No’ and ‘Unknown’. We use the similar strategy to define patient’s death status where 1 indicates the death and 0 indicates alive which includes alive and unknown.

### Statistical analysis

Descriptive statistics were used to characterize the disease severity and clinical outcomes for COVID-19 cases. We used Chi-square test to compare differences between groups. We used multinomial logistic and logistic regression models to explore the association between socio- demographic characteristics and symptom severity, hospitalization and death in COVID-19 cases. We reported odds ratios (OR) and 95% confidence intervals (95% CI) for each model in tables and forest plots. P-value <0.05 were considered statistically significant. All statistical analysis were performed using SAS software version 9.4 (SAS Institute, Inc., Cary, NC) and R software (version 3.6.2).

## Results

### Demographics and disease severity

A total of 280,177 COVID-19 cases were included in this analysis, 58.4% were aged 18-49 years old (mean: 45.7; SD: 18.9), 54.9% were female, 53.7% were White, 6.5% were Hispanic or Latino and the majority were from metropolitan area (81.7%) (higher than the percent of the overall population in SC lives in metropolitan areas: 58.4% in 2016). Slightly over half (51.5% or 144,157/280,177) of the cases were asymptomatic, 34.4% (96,252/280,177) had mild symptom, 14.2% (39,768/280,177) had moderate/severe symptom. Figure 1 illustrates the frequency of symptoms, with cough, headache, myalgia, taste loss, and fever as the most common ones. Demographic characteristics by disease severity is shown in Table 1. Compared to asymptomatic cases, individuals ≥65 years of age, females, White, Hispanic or Latino, and individuals living in small towns were more likely to experience moderate/severe symptoms. (Table 1)

**Table 1.**
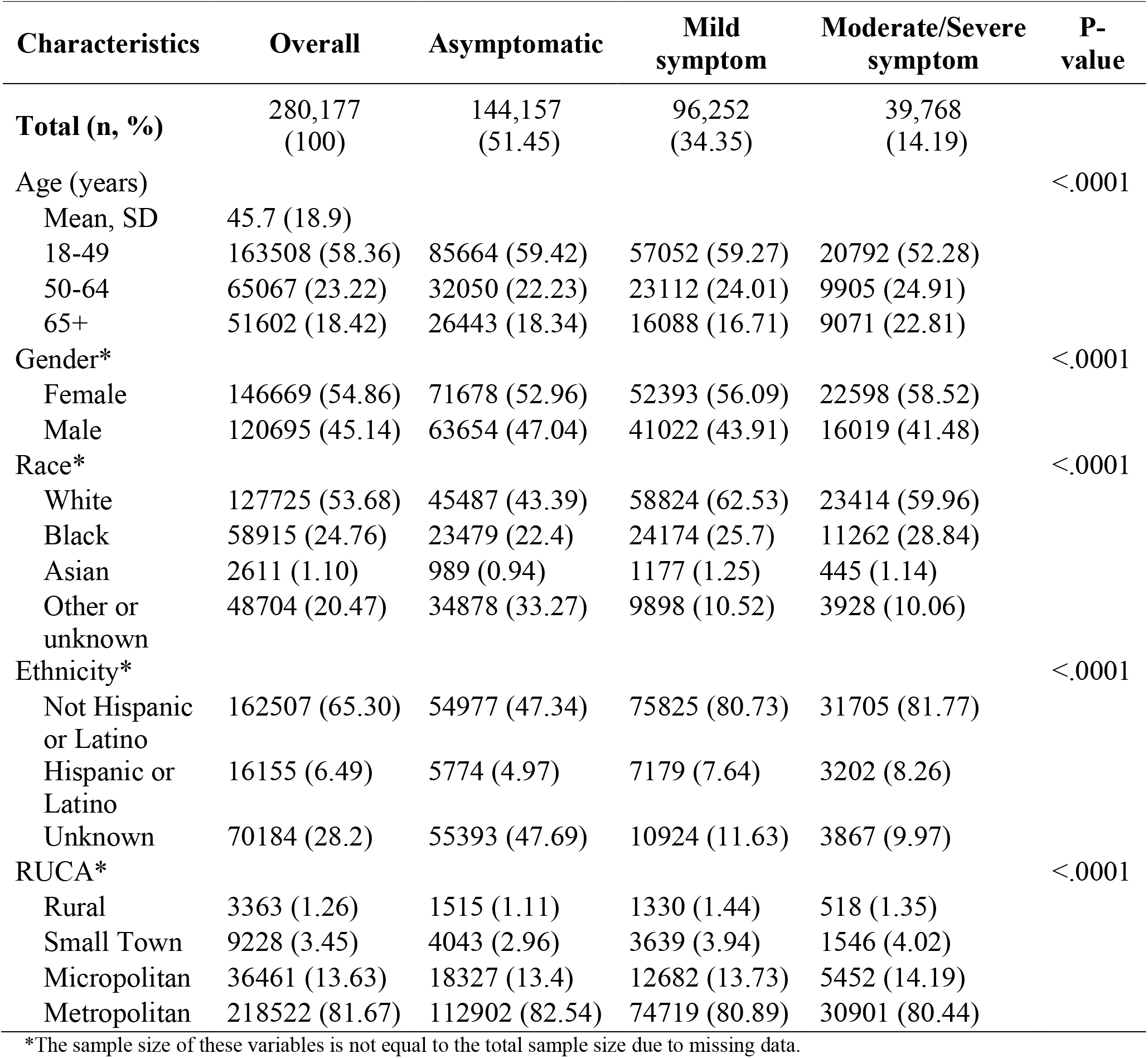
Socio-demographic distribution of COVID-19 symptom severity.

| Characteristics | Overall | Asymptomatic | Mild symptom | Moderate/Severe symptom | P-value |
| --- | --- | --- | --- | --- | --- |
| **Total (n, %)** | 280,177<br>(100) | 144,157<br>(51.45) | 96,252<br>(34.35) | 39,768<br>(14.19) |  |
| Age (years) |  |  |  |  | <.0001 |
| Mean, SD | 45.7 (18.9) |  |  |  |  |
| 18-49 | 163508 (58.36) | 85664 (59.42) | 57052 (59.27) | 20792 (52.28) |  |
| 50-64 | 65067 (23.22) | 32050 (22.23) | 23112 (24.01) | 9905 (24.91) |  |
| 65+ | 51602 (18.42) | 26443 (18.34) | 16088 (16.71) | 9071 (22.81) |  |
| Gender* |  |  |  |  | <.0001 |
| Female | 146669 (54.86) | 71678 (52.96) | 52393 (56.09) | 22598 (58.52) |  |
| Male | 120695 (45.14) | 63654 (47.04) | 41022 (43.91) | 16019 (41.48) |  |
| Race* |  |  |  |  | <.0001 |
| White | 127725 (53.68) | 45487 (43.39) | 58824 (62.53) | 23414 (59.96) |  |
| Black | 58915 (24.76) | 23479 (22.4) | 24174 (25.7) | 11262 (28.84) |  |
| Asian | 2611 (1.10) | 989 (0.94) | 1177 (1.25) | 445 (1.14) |  |
| Other or unknown | 48704 (20.47) | 34878 (33.27) | 9898 (10.52) | 3928 (10.06) |  |
| Ethnicity* |  |  |  |  | <.0001 |
| Not Hispanic or Latino | 162507 (65.30) | 54977 (47.34) | 75825 (80.73) | 31705 (81.77) |  |
| Hispanic or Latino | 16155 (6.49) | 5774 (4.97) | 7179 (7.64) | 3202 (8.26) |  |
| Unknown | 70184 (28.2) | 55393 (47.69) | 10924 (11.63) | 3867 (9.97) |  |
| RUCA* |  |  |  |  | <.0001 |
| Rural | 3363 (1.26) | 1515 (1.11) | 1330 (1.44) | 518 (1.35) |  |
| Small Town | 9228 (3.45) | 4043 (2.96) | 3639 (3.94) | 1546 (4.02) |  |
| Micropolitan | 36461 (13.63) | 18327 (13.4) | 12682 (13.73) | 5452 (14.19) |  |
| Metropolitan | 218522 (81.67) | 112902 (82.54) | 74719 (80.89) | 30901 (80.44) |  |

\*The sample size of these variables is not equal to the total sample size due to missing data.

### Hospitalization and mortality

Hospitalization occurred in 5.2% (n=14,451) of the COVID-19 cases and 1.9% (n=5,308) died from COVID-19. Individuals who were male, older, non-White race, residing in the small-town areas, and having more moderate/severe symptoms were more likely to be hospitalized or die (Table 2). Individuals with moderate/severe symptoms accounted for the largest proportion of all the clinical outcomes (i.e., hospitalization [3.0%], ICU admission [0.6%], respiratory support [1.7%], and death [0.9%]). (Figure 2)

**Table 2.**
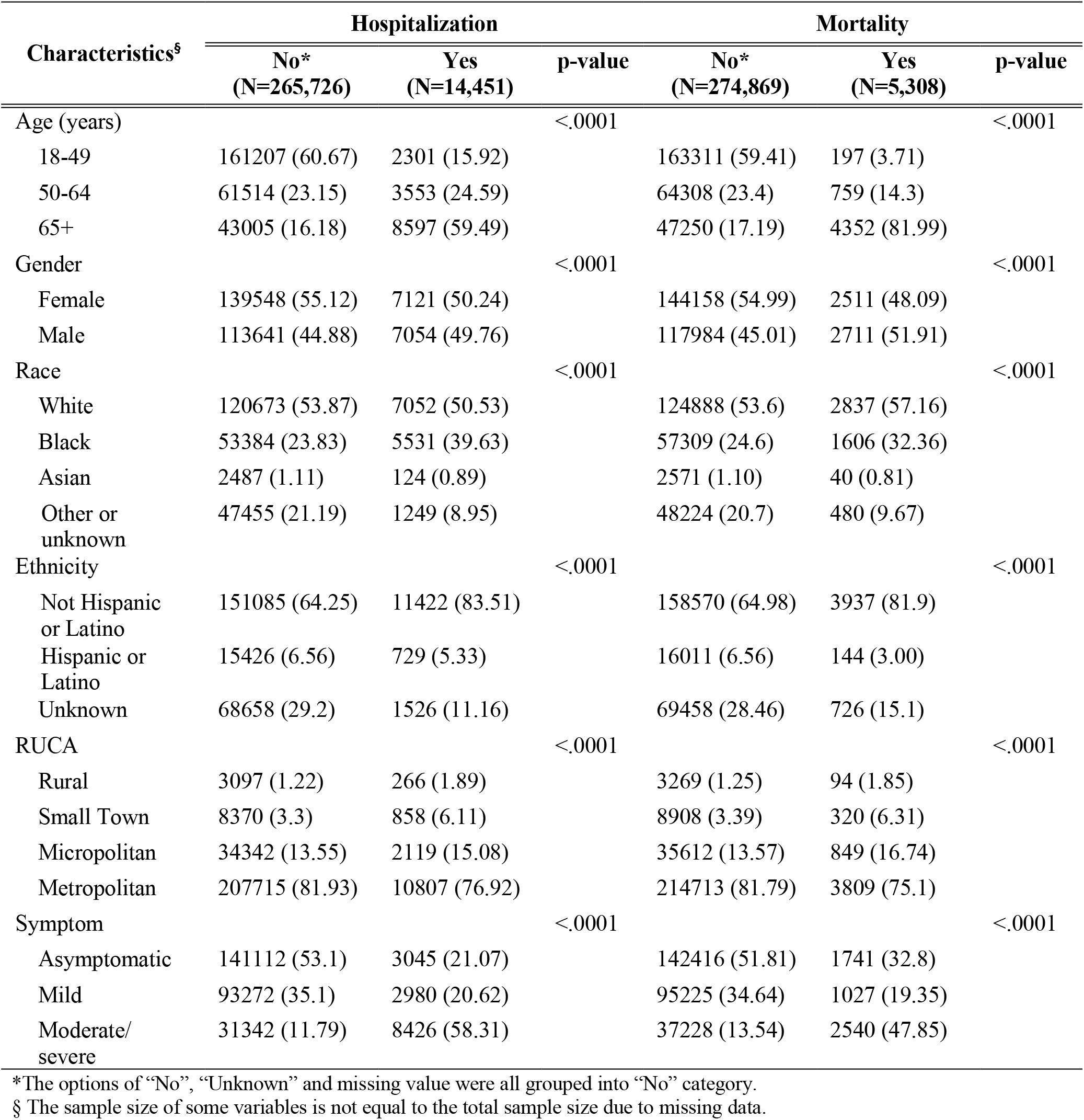
Socio-demographic disparities of COVID-19 hospitalization and mortality (n=280,177)

| Characteristics<sup>§</sup> | Hospitalization |  |  | Mortality |  |  |
| --- | --- | --- | --- | --- | --- | --- |
|  | No*<br>(N=265,726) | Yes<br>(N=14,451) | p-value | No*<br>(N=274,869) | Yes<br>(N=5,308) | p-value |
| Age (years) |  |  | <.0001 |  |  | <.0001 |
| 18-49 | 161207 (60.67) | 2301 (15.92) |  | 163311 (59.41) | 197 (3.71) |  |
| 50-64 | 61514 (23.15) | 3553 (24.59) |  | 64308 (23.4) | 759 (14.3) |  |
| 65+ | 43005 (16.18) | 8597 (59.49) |  | 47250 (17.19) | 4352 (81.99) |  |
| Gender |  |  | <.0001 |  |  | <.0001 |
| Female | 139548 (55.12) | 7121 (50.24) |  | 144158 (54.99) | 2511 (48.09) |  |
| Male | 113641 (44.88) | 7054 (49.76) |  | 117984 (45.01) | 2711 (51.91) |  |
| Race |  |  | <.0001 |  |  | <.0001 |
| White | 120673 (53.87) | 7052 (50.53) |  | 124888 (53.6) | 2837 (57.16) |  |
| Black | 53384 (23.83) | 5531 (39.63) |  | 57309 (24.6) | 1606 (32.36) |  |
| Asian | 2487 (1.11) | 124 (0.89) |  | 2571 (1.10) | 40 (0.81) |  |
| Other or unknown | 47455 (21.19) | 1249 (8.95) |  | 48224 (20.7) | 480 (9.67) |  |
| Ethnicity |  |  | <.0001 |  |  | <.0001 |
| Not Hispanic or Latino | 151085 (64.25) | 11422 (83.51) |  | 158570 (64.98) | 3937 (81.9) |  |
| Hispanic or Latino | 15426 (6.56) | 729 (5.33) |  | 16011 (6.56) | 144 (3.00) |  |
| Unknown | 68658 (29.2) | 1526 (11.16) |  | 69458 (28.46) | 726 (15.1) |  |
| RUCA |  |  | <.0001 |  |  | <.0001 |
| Rural | 3097 (1.22) | 266 (1.89) |  | 3269 (1.25) | 94 (1.85) |  |
| Small Town | 8370 (3.3) | 858 (6.11) |  | 8908 (3.39) | 320 (6.31) |  |
| Micropolitan | 34342 (13.55) | 2119 (15.08) |  | 35612 (13.57) | 849 (16.74) |  |
| Metropolitan | 207715 (81.93) | 10807 (76.92) |  | 214713 (81.79) | 3809 (75.1) |  |
| Symptom |  |  | <.0001 |  |  | <.0001 |
| Asymptomatic | 141112 (53.1) | 3045 (21.07) |  | 142416 (51.81) | 1741 (32.8) |  |
| Mild | 93272 (35.1) | 2980 (20.62) |  | 95225 (34.64) | 1027 (19.35) |  |
| Moderate/severe | 31342 (11.79) | 8426 (58.31) |  | 37228 (13.54) | 2540 (47.85) |  |

\*The options of “No”, “Unknown” and missing value were all grouped into “No” category.
§ The sample size of some variables is not equal to the total sample size due to missing data.

**Figure 2.**
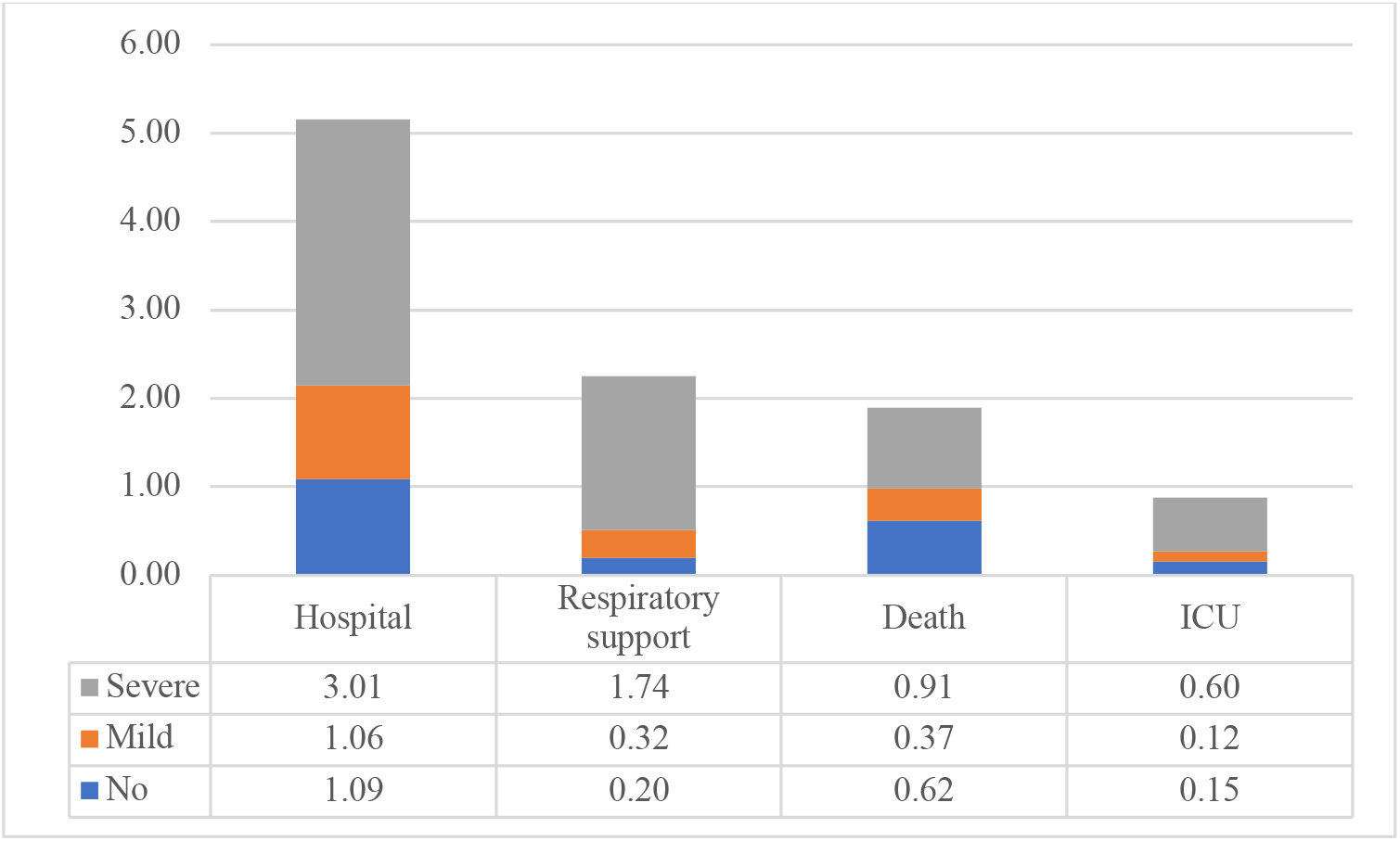
Clinical outcomes of COVID-19 patients stratified by illness severity (n=280,177)

### Association between socio-demographics, symptom severity, hospitalization and mortality

Older individuals were more likely to experience moderate/severe symptoms, require hospitalization and die from COVID-19. Compared to males, females were more likely to have mild or moderate/severe symptoms, but males were more likely to require hospitalization and die. (Table 3 &4 & Supplement Figure 1)

**Table 3.** Associations between socio-demographics and COVID-19 disease severity.

| **Characteristics** | **Mild vs**<br>**Asymptomatic**<br>**(OR, 95%CI)** | **P-value** | **Moderate/Severe vs**<br>**Asymptomatic**<br>**(OR, 95%CI)** | **P-value** |
| --- | --- | --- | --- | --- |
| Age (years) |  |  |  |  |
| 18-49 | Ref |  |  |  |
| 50-64 | 0.98 (0.96,1.01) | 0.1639 | 1.16 (1.13,1.20) | <0.0001 |
| 65+ | 0.73 (0.71,0.75) | <0.0001 | 1.14 (1.11,1.18) | <0.0001 |
| Gender |  |  |  |  |
| Female | Ref |  |  |  |
| Male | 0.90 (0.89,0.92) | <0.0001 | 0.82 (0.80,0.84) | <0.0001 |
| Missing | 0.88 (0.84,0.93) | <0.0001 | 0.85 (0.80,0.92) | <0.0001 |
| Race |  |  |  |  |
| White | Ref |  |  |  |
| Black | 0.83 (0.82,0.85) | <0.0001 | 1.00 (0.97,1.03) | 0.7878 |
| Asian | 0.87 (0.79,0.95) | 0.0017 | 0.87 (0.77,0.97) | 0.0143 |
| Other or<br>Unknown | 0.31 (0.30,0.32) | <0.0001 | 0.32 (0.31,0.34) | <0.0001 |
| Missing | 0.11 (0.10,0.11) | <0.0001 | 0.09 (0.09,0.10) | <0.0001 |
| Ethnicity |  |  |  |  |
| Not Hispanic<br>or Latino | Ref |  |  |  |
| Hispanic or<br>Latino | 1.74 (1.67,1.81) | <0.0001 | 2.07 (1.96,2.18) | <0.0001 |
| Unknown | 0.30 (0.29,0.30) | <0.0001 | 0.26 (0.25,0.27) | <0.0001 |
| Missing | 0.14 (0.13,0.14) | <0.0001 | 0.14 (0.14,0.15) | <0.0001 |
| RUCA |  |  |  |  |
| Metropolitan | Ref |  |  |  |
| Rural | 1.19 (1.10,1.30) | <0.0001 | 1.06 (0.95,1.18) | 0.3183 |
| Small Town | 1.21 (1.15,1.28) | <0.0001 | 1.15 (1.08,1.23) | <0.0001 |
| Micropolitan | 0.98 (0.96,1.01) | 0.2243 | 0.98 (0.95,1.02) | 0.3505 |
| Missing | 1.00 (0.95,1.05) | 0.9867 | 0.88 (0.82,0.94) | <0.0001 |

There were several differences in symptom presentation by race as seen in Table 3. Notably, Hispanic or Latino had higher odds to report both mild and moderate/severe symptoms than its counterpart. For hospitalization, Asian, Black, and Hispanics all had higher odds of hospital admission. Only Blacks had higher odds of death comparing with Whites, no significant mortality difference was detected for Asian and Hispanics. (Table 4 & Supplement Figure 1)

**Table 4.** Associations between socio-demographics and COVID-19 hospitalization and mortality.

| Characteristics | Hospitalization |  | Mortality |  |
| --- | --- | --- | --- | --- |
|  | (OR, 95%CI) | p-value | (OR, 95%CI) | p-value |
| Age (years) |  |  |  |  |
| 18-49 | Ref |  | Ref |  |
| 50-64 | 3.97 (3.76,4.20) | <0.0001 | 9.07 (7.75,10.62) | <0.0001 |
| 65+ | 16.83 (15.99,17.72) | <0.0001 | 71.04 (61.5,82.06) | <0.0001 |
| Gender |  |  |  |  |
| Female | Ref |  |  |  |
| Male | 1.37 (1.32,1.43) | <0.0001 | 1.36 (1.28,1.44) | <0.0001 |
| Missing | 0.63 (0.56,0.73) | <0.0001 | 0.52 (0.42,0.65) | <0.0001 |
| Race |  |  |  |  |
| White | Ref |  |  |  |
| Black | 2.39 (2.29,2.5) | <0.0001 | 1.46 (1.36,1.56) | <0.0001 |
| Asian | 1.37 (1.12,1.68) | 0.0022 | 1.28 (0.92,1.79) | 0.1434 |
| Other or Unknown | 1.11 (1.03,1.20) | 0.008 | 1.01 (0.9,1.14) | 0.8561 |
| Missing | 0.65 (0.58,0.72) | <0.0001 | 0.68 (0.59,0.78) | <0.0001 |
| Ethnicity |  |  |  |  |
| Not Hispanic or Latino | Ref |  |  |  |
| Hispanic or Latino | 1.42 (1.29,1.56) | <0.0001 | 1.03 (0.85,1.23) | 0.7776 |
| Unknown | 0.66 (0.62,0.70) | <0.0001 | 0.81 (0.74,0.9) | <0.0001 |
| Missing | 0.95 (0.87,1.04) | 0.2791 | 1.37 (1.22,1.54) | <0.0001 |
| RUCA |  |  |  |  |
| Metropolitan | Ref |  |  |  |
| Rural | 1.19 (1.03,1.38) | 0.0213 | 1.19 (0.95,1.49) | 0.1285 |
| Small Town | 1.30 (1.19,1.42) | <0.0001 | 1.38 (1.21,1.56) | <0.0001 |
| Micropolitan | 0.96 (0.91,1.02) | 0.1702 | 1.11 (1.02,1.2) | 0.0135 |
| Missing | 0.87 (0.78,0.97) | 0.0139 | 1.49 (1.29,1.71) | <0.0001 |
| Symptom |  |  |  |  |
| Asymptomatic | Ref |  |  |  |
| Mild | 1.29 (1.22,1.37) | <0.0001 | 0.87 (0.8,0.95) | 0.0017 |
| Severe | 11.61 (11.04,12.21) | <0.0001 | 4.72 (4.39,5.07) | <0.0001 |

The symptom presentation varied between different residence types as well (Table 3). Comparing with individuals living in the metropolitan area, those living in the small towns were more likely to exhibit mild and moderate/severe symptoms. When comparing the hospitalization rates, patients living in the rural area and small towns were more likely to be hospitalized. No significant difference was detected for mortality between patients living in the rural area and metropolitan area but living in small towns was positively associated with mortality. (Table 3 &4 & Supplement Figure 1)

## Discussion

To the best of our knowledge, this is the first statewide population-based US study of COVID-19 case investigating symptom manifestations and disease progression. Most existing studies were either based on non-representative samples or underrepresented the outpatient COVID-19 cases. For example, US national study-COVID-19-Associated Hospitalization Surveillance Network (COVID-NET) conducted COVID-19 related research on hospitalized COVID-19 patients(18). Few studies have systematically collected data on COVID-19 patients from the statewide case report form that is representative of both inpatient and outpatient COVID-19 cases. In addition, the standardized case report form in our study measured various symptoms including asymptomatic or mild symptoms. Our population-based study overcomes the sampling selection bias in previous studies and provided added rigor to describe the presenting symptoms and clinical outcomes of COVID-19 patients.

Consistent with previous studies in both US(23) and China(24), the study found common symptoms to include as cough, headache, fever, and myalgia, while gastrointestinal symptoms (e.g., nausea or vomiting and diarrhea) were uncommon. Importantly, loss of taste or smell, a prominent symptom of SARS-CoV-2 infection (25), was observed in a large proportion of COVID-19 patients in our study (20%). This could be indicative of neurological issues and needs future monitoring.

Studies show that asymptomatic COVID-19 infected persons play a significant role in active transmission. According to the CDC’s best estimate, the infectiousness of asymptomatic individuals relative to symptomatic is 75%(26). Thus, asymptomatic cases “substantially contribute to community transmission, making up at least 50% of the driving force” of COVID- 19 infections(4). However, determining the actual number of asymptomatic COVID-19 cases has been a significant challenge for researchers and public health officials. The proportion of asymptomatic cases in the present study is comparable with a narrative review(4) but higher than the best estimate (30%) reported by the CDC(26). The data sources in our study incorporated statewide data about daily testing capacity and changes in testing rates over time, so we believe the findings from this study could provide a more accurate estimation of the proportion of COVID-19 infections that are asymptomatic. Given the large proportion of asymptomatic cases, it is crucial that everyone including individuals who do not show symptoms-adhere to public health guidelines, such as mask wearing and social distancing.

Consistent with previous studies, older age was associated with hospitalization and mortality.(7, 18, 27) Older patients were less likely to display mild symptoms but more moderate/severe symptoms of COVID-19. Others have reported that older individuals with COVID-19 often present with non-specific and atypically symptoms such as delirium, postural instability or diarrhea(28-30), rather than typical respiratory symptoms and fever. According to a meta-analysis, gastrointestinal symptoms including vomiting and diarrhea are strong predictors of developing severe COVID-19 illness(31). Some atypical symptoms (e.g., delirium) occurred in older people might not be measured in our study. That may partially explain why older people with fewer common symptoms are not necessarily less likely to develop severe symptoms. We need to prioritize the needs of older people in the response to the COVID-19 pandemic.

While there is not a significant difference in the proportion of male and female COVID-19 cases, a disparity in hospitalization and death was observed. According to a recent meta-analysis using data from 46 countries and 44 US states, the gender differences observed in COVID-19 is a worldwide phenomenon with men being more likely to die or require intensive care unit (ICU) admission for COVID-19(32). The driving factors behind the gender difference may include the fundamental differences in the immune response (e.g., CD4+ T cells, type 1 interferon, estrogen level),(33, 34) cultural and behavioral differences (e.g., smoking, handwashing frequency)(35, 36). An appreciation of how gender influences COVID-19 outcomes will have important implications for clinical management and mitigation strategies for this disease.

The COVID-19 pandemic has highlighted persistent racial-/ethnic- health disparities in the US. Black, Asian, and Minority Ethnic groups (termed as ‘BAME’ in the UK(37)) are overrepresented among cases of hospitalization and deaths from COVID-19 in the present study. This is consistent with findings from both national and international studies(8, 38). The likely causes of racial-/ethnic- health disparities in COVID-19 outcomes were discussed extensively in prior studies(9, 11, 14, 16, 37, 39-44). Some literature argued that minority communities may be more susceptible to severe complications of COVID-19 because of existing disparities in underlying conditions known to be associated with COVID-19 mortality, including hypertension, cardiovascular disease, kidney disease, and diabetes. However, studies conducted in New York and Louisiana published in *JAMA* and *NEJM*, found that once hospitalized, Black patients were less likely than White patients to develop critical illness or die after adjusting for comorbidity and neighborhood characteristics(9, 11). The structural racism, which shapes the distribution of social determinants of health and social risk factors, such as poverty, healthcare access, service occupation, are associated with the COVID-19 pandemic(40). These factors and others are interrelated and influence a wide range of health and quality of life outcomes and risks, including SARS-CoV-2 infection, hospitalizations, and deaths in areas where racial and ethnic minority groups live, learn, work, play, and worship(9, 19, 45, 46). This supports the assertion that existing structural determinants (e.g., housing, economic stability, and work circumstances) pervasive in Black and Hispanic communities may explain the disproportionately higher out-of-hospital deaths due to COVID-19 infections in these populations. It might indicate that black persons are less likely to be identified in the outpatient setting, potentially reflecting differences in health care access or utilization. The percentage of presenting symptoms among Black people, which is lower than White in our study, might also be underreported due to lack of access to healthcare(40). For Asians, the higher expression of ACE2 (the receptor for SARS-CoV-2), which is more predominant in Asian men(47), might explain the higher susceptibility to COVID- 19 in this subgroup. Additional research is needed to more fully understand the impact of race- /ethnic- disparities on COVID-19 outcomes. Given the overrepresentation of race-/ethnic-minority patients with critical outcomes within this cohort, it is important for public health officials to ensure that prevention activities prioritize communities and racial/ethnic groups most affected by COVID-19.

The findings in this report are subject to several limitations. First, there were missing values for the outcomes, such as hospitalization and mortality. The missing values might cause inaccurate estimation in clinical outcomes. For example, if more hospitalized or dead patients were misclassified into missing data, the outcomes might be underestimated in this study. Second, some important variables, such as underlying conditions, were not included in the current analysis as these conditions would merit separate analyses because of their clinical significance to COVID-19 research.

Despite of the limitations, this study is still one of the first US statewide population-based study using the entire population to investigate the presenting symptoms and clinical outcomes of COVID-19 patients. Such a population-based study can minimize sampling selection bias and is more representative of all COVID-19 cases. Our results revealed that severe illness was strongly associated with hospitalization and mortality. However, the differences in the symptom distribution are not reflected in disparities in hospitalization and mortality in certain gender- and racial- minority groups. The findings from this study reinforce the fact that underlying health system disparities remain a challenge. South Carolina is often reflective of the “Deep South” states. Preexisting structural disparities were exacerbated during COVID-19 and put the already vulnerable populations at more risk. Rural residence, racial and ethnic social determinants of health unfortunately remain predictors of poor health outcomes for COVID-19 patients. The effects are ongoing, making it a priority for interventions and policies to alleviate these problems in both the short and long-term.

## Data Availability

Given the confidentiality and privacy concerns of patient's information, the data used in this study was not publicly available.

