## Supplement Figure 1 for "Demographic disparities in clinical outcomes of COVID-19: data from a statewide cohort in South Carolina"

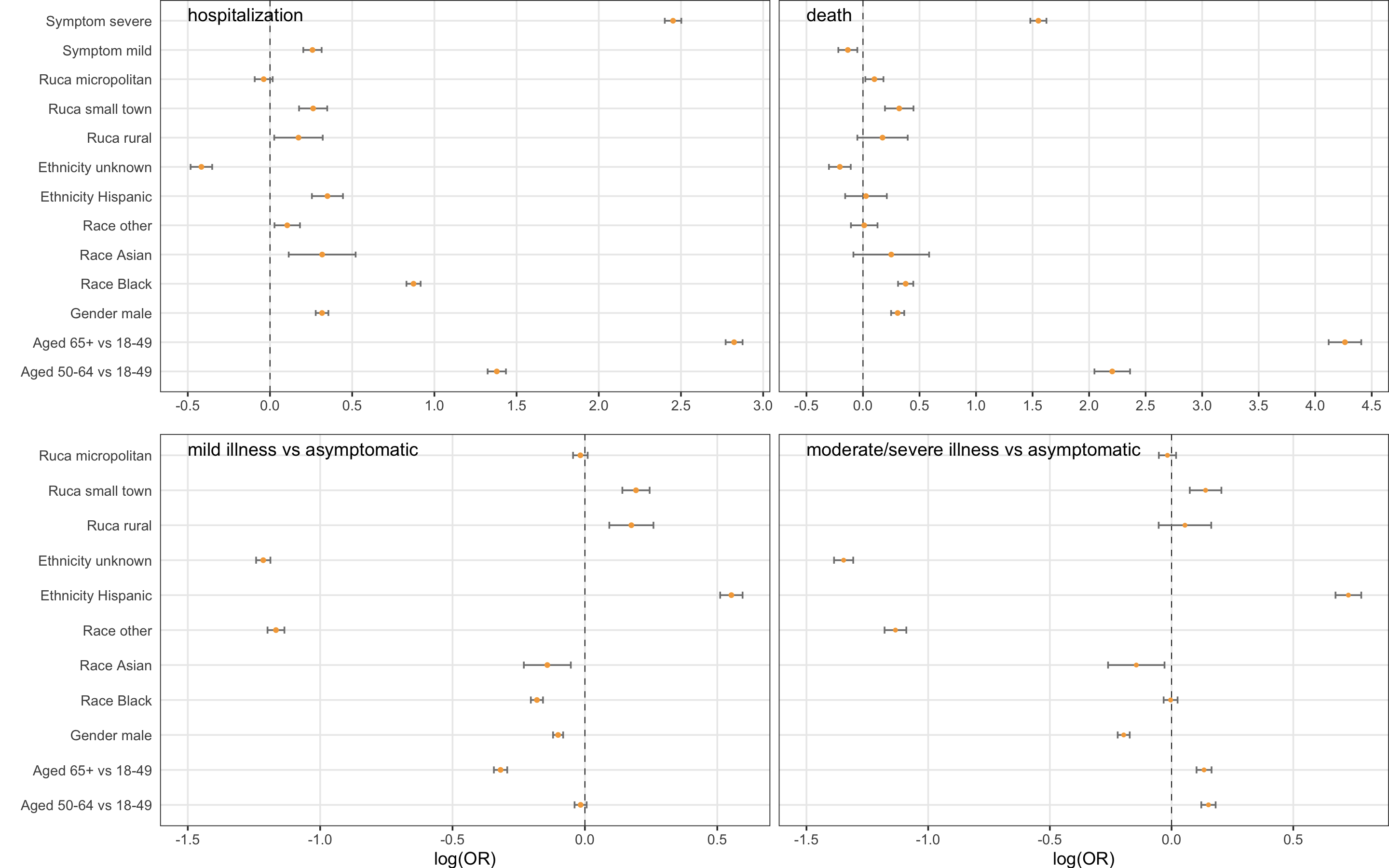

**Figure 1 Log (Odds Ratios) and 95% CI for illness severity, hospitalization, and mortality among statewide COVID-19 patients (n=280,177) in South Carolina, March 4, 2020-December 31, 2020**
